# The Relationship between Chronological Age, Dental Age, and Salivary Alkaline Phosphatase in Indonesian Children Aged 8-14 Years: A Cross-Sectional Study

**DOI:** 10.1101/2021.01.14.21249792

**Authors:** Sindy Cornelia Nelwan, Soegeng Wahluyo, Firli Cahaya Khairani, Ricardo Adrian Nugraha, Haryono Utomo, Udijanto Tedjosasongko

**Affiliations:** Department of Pediatric Dentistry, Faculty of Dental Medicine, Universitas Airlangga; Department of Cardiology and Vascular Medicine, Faculty of Medicine, Universitas Airlangga; Department of Forensic Odontology, Faculty of Dental Medicine, Universitas Airlangga

**Keywords:** alkaline phosphatase, biological age, chronological age, dental age, saliva

## Abstract

**Background:** Inability to predict dental age accurately has long been the Achilles heel of pediatric dentistry. Whilst dental age has an important aspect in clinical practice, saliva can be one of the most practically important diagnostic tools to estimate biological age.

**Aims:** This study was aimed to analyze a correlation between chronological age, dental age, and salivary alkaline phosphatase in Indonesian children aged 8-14 years.

**Methods:** This study was an observational study with cross-sectional design. Twenty healthy children (10 boys; age 10.504 ± 1.895 years) were selected by a consecutive sampling. Chronological age was assessed by recording date of birth. Dental age was assessed by orthopantamogram following Demirjian’s method. Salivary samples were collected by passive drool method and estimation of alkaline phosphatase was done by autoanalyzer.

**Results:** Normality test was assessed using Kolmogorov-Smirnov and Shapiro-Wilk test. Statistical analyses were assessed using Spearman’s rank-order correlation coefficients. Results are presented as mean ± standard deviation (SD). Mean chronological age was10.504 ± 1.895 years, mean Demirjian’s score was 91.74 ± 5.972, and mean salivary alkaline phosphatase was 291.563 ± 115.135 pg/ml. There is a very strong positive correlation between chronological age and dental age (r=+0.804; *p*<0.001). On the contrary, levels of salivary alkaline phosphatase was negatively correlated to dental age (r=-0.780; *p*<0.001) and chronological age (r=-0.508; *p*=0.022).

**Conclusions:** This study showed a strong correlation between dental age, chronological age, and salivary alkaline phosphatase; hence, estimation of dental age and salivary alkaline phosphatase in Indonesian children during early and late childhood has significant meaning to chronological age.

## INTRODUCTION

Age estimation has gained increasing importance in many practical decisions, such as legal medicine, forensic science, anthropology and clinical practice^1^. In legal medicine, age estimation may help judges, courts and government to determine the exact age of persons whose actual age is unknown, in accordance with the rule of ethical, legal, and criminal perspectives^2^. In forensic science, age estimation can guide investigators to the correct identity of unknown bodies among a large number of possible matches and in the identification, work associated with mass disasters^3^. In anthropology, predicting an age could be a way to establish the identity of human remains^4-5^. In clinical practice, clinician shouldn’t only rely on chronological age for objective health indicators, subjective health perceptions, and psychological well-being indicators, because there are some conditions or development variations making chronological age and biological age might not be the same^6-7^.

In instances where chronological age is unknown, unclear, undocumented or missing, there are several ways to predict biological age as accurate as chronological age^8^. At present, biological age can be measured by bone age and dental age^9^. Bone age and dental age have been assessed to determine to what extent they are correlated for diagnostic purposes^9^. Measurement of bone age is based on skeletal indicators such as hand-wrist bone ossification, changes in pubic symphysis, and fusion of cranial sutures which are utilized for to indicate level of biological and structural maturity better than the chronological age^10^. Measurement of dental age is based on morphologic methods, biochemistry methods, and radiographic methods which show dental maturation^11^. Considering that states of dental mineralization are much less affected by nutritional^12^ and hormonal^13^ variations than states of bone mineralization, dental age provides more reliable estimation of biological age than bone age^14^.

Long time ago, researchers and clinicians have discovered tooth development that takes place from early fetal life to approximately 20 years of age^15^. As long as dental age estimation is concerned, several methods have been proposed for assessing dental development, which is generally referred to as dental age^16^. Morphologic and radiographic methods (Cameriere’s, Schour-Massler’s, Demirjian’s, and Kvaal’s method) are useful in living humans^17^, whereas biochemistry methods (Gustafson’s and Johanson’s method, Bang and Ramm method, aspartic acid racemization and cemental annulation technique) are useful in dead victims^18^.

Demirjian’s method has been the most widely used method and considered as the gold standard for dental age estimation^19^. Several literatures supports Demirjian’s method for dental age estimation in Northeastern Turkish children^20^, Malay population^21^, Western Chinese children^22^, Malaysian children^23^, Belgaum population^24^, Lucknow children^25^, Indian children^19^, and Indonesian children^26-28^.

Some alternative strategies such as biochemistry parameters will increase the precision and reliability of dental age estimation. In this decade, interest in saliva as a diagnostic medium to estimate children’s dental age has increased exponentially. Salivary components such as enzymes, immunoglobulins, inorganic materials and ions have different concentration in the different age^29^. From some salivary components, there are few components that sounds interesting to be observed in accordance to dental aging and maturation. One of them is alkaline phosphatase, in which the application for bone growth isn’t no longer debatable^30^.

Alkaline phosphatase (ALP) itself is a hydrolase intracellular enzyme participating in the remineralization processes of enamel^31^. ALP is a membrane-bound glycoprotein found on growing cells and most cell membranes in the body and physiologically occurs during bone formation in developmental stages^32^. ALP is synthesized by the osteoblasts and is presumed to be involved in the calcification of bone matrix and bone mineralization, varying from 77% to 89% in children and from 58% to 67% in adults^33^. Since serum ALP concentrations have limitations in their great discrepancies within age and sex^34^, we try to estimate salivary ALP concentrations which tend to be more stable and could represent true age with good approximation.

## AIMS OR OBJECTIVES

This study aimed to analyze a correlation between chronological age, dental age, and salivary alkaline phosphatase in children aged 8-14 years. The authors proposed the following specific objectives: 1) to analyze the predictive capacities of Demirjian method for age estimation in an Indonesian children population; 2) to detect ALP levels in saliva and to correlate it with the dental age and to project it as a noninvasive tool for assessment of chronological age; 3) to compare and correlate the magnitude of agreement of the dental age and salivary ALP to construct a global model with a predictive value of chronological age; 4) to carry out empirical testing of the model’s adjustment, comparing real values with predicted values.

## METHODS

### Study design and study setting

An analytic observational study with cross-sectional study design was done in this study. This cross-sectional study involved analysis of confounding variables. Chronological age, dental age and salivary ALP concentration were included as the major variables and sex as the confounding variables. This study was conducted in the Department of Pediatric Dentistry, Faculty of Dental Medicine, Universitas Airlangga, during 1-31 January 2018. Dental examination and radiographs were taken at Universitas Airlangga Oral and Dental Hospital. Samples of saliva were brought and analyzed at Institute of Tropical Diseases - Universitas Airlangga. We observed and analyzed data from subjects with inclusion and exclusion criteria as follow. The findings of the three variables were compared between the study participants.

### Subject population

Indonesian healthy children of Javanese ethnicity who were treated at the Universitas Airlangga Oral and Dental Hospital, Surabaya during 1-31 January 2018, included girls and boys at the same proportion. A consecutive sampling was done in this study to achieve 24 subjects to participate with the following inclusion and exclusion criteria.

### Inclusion criteria

- Healthy children aged 8-14 years recruited from the community
- Understand and able to cooperate to the research protocol
- Subjects’ parents or legal representatives have signed the written consent in accordance with our institutional policies

### Exclusion criteria

- History of musculoskeletal conditions such as rickets or recent fractures
- History of maxillofacial trauma with significant numbers of missing teeth
- History of childhood obesity, cancer, and type I diabetes mellitus
- History of liver or bile ducts diseases, anemia, or hyperparathyroidism
- History of gingivitis, dental caries, severe bacterial infection within 12 months
- History of maternal alcohol use during pregnancy and/or breast-feeding
- Orthopantomogram distortion due to incomplete exposure, subject movement during exposure, or improper positioning of subject
- Subjects who are unable to perform the test or uncooperate

### Method to calculate chronological age

Subjects’ birth dates were noted after analyzing their specific identity proofs. Consistent with recent age form consensus definitions from Merriam-Webster, chronological age or calendar age was measured of an individual’s age based on the calendar date on which he or she was born^35^.

### Method to estimate dental age

Subjects’ dental age were analyzed with Orthopantomograph OP200 D Dental Digital Panoramic X-ray Unitat 71-73 V and 10 mA. Exposure time was 13-14 seconds with 4096 bits grey scale level. Subjects’ radiographs selection criteria were as follows:

- Only high-quality panoramic radiographs, with respect to angulations, contrast and correct positioning, were included in this study.
- Radiographs should be free from any artifacts.
- Radiographs should not show any developmental anomalies of teeth related to size, shape and structure of teeth.

Estimation of dental age was done using Demirjian′s method considering 7 permanent left mandibular teeth from central incisor to II molar^36^. Determination of dental age was based upon the rate of development and calcification of tooth buds. The developmental stage of each tooth was assessed and then each tooth was given a score according to its stage of development using the score table. Adding 7 individual scores from permanent central incisor to 2^nd^ permanent molar gives a maturity score, maturity score will be converted into dental age using conversion chart^37^.

Demirjian’s tooth mineralization stages are as follows:

Stage A: Beginning mineralization of separate cusps.

Stage B: Fusion of cusps.

Stage C: Beginning of dentinal deposits is seen.

Stage D: Crown formation completed down to the cemento-enamel junction.

Stage E: The root length is less than the crown height.

Stage F: The root length is equal to or greater than crown height.

Stage G: The walls of the root canal are parallel and its apical end is still partially open.

Stage H: The apical foramen is completed. [36]

In order to assess the reproducibility of our analysis, a subset of 24 orthopantomogram radiographs included in this study were randomly and blindly chosen to be reviewed by 4 independent observers within a period of 2 weeks. Independent observers compared the radiographic and actual tooth eruptions. Results were calculated from the average of each independent observers’ score.

### Method to estimate salivary alkaline phosphatase concentration

A volume of 1.5 ml of unstimulated saliva samples were collected in sample container from 20 subjects. Subjects were instructed not to take food for 2 hours prior to saliva collection. All subjects were asked to collect saliva into a sample container. Samples of saliva was brought and analyzed at Institute of Tropical Diseases, Universitas Airlangga. Sample was collected and centrifuged at 3000 rpm for 15 minutes. The reagents used in this analysis were p-Nitrophenylphospate, and 2-amino-2-methyl-1-propanol. Using Bio-Rad iMark Microplate Absorbance Reader, ALP was estimated and the value of ALP was expressed in pg/ml. The readings obtained on the screen of analyzer were noted.

### Statistical analysis

All the data was recorded, tabulated and statistically analyzed using the Statistical Package for Social Sciences (SPSS, version 13.0; SPSS Inc., Chicago, IL) software. Statistical presentation and analysis of the present study was conducted using mean ± standard derivative (SD). The normality criterion was evaluated using Kolmogorov-Smirnov and Saphiro–Wilk test. The quantitative variables were compared using the Spearman correlation test. The statistical differences were significant if *p*-values < 0.05.

## RESULT

A total 24 subjects were selected who matched inclusion and exclusion criteria in the study, of which 20 subjects were included in the study. From 24 subjects selected to participate in this study, 4 subjects were excluded as follows: 1 had fear of dental x-ray for panoramic scanning, 2 failed to collect saliva samples, and 1 parent rejected to participate after information to consent had been given due to lack of time.

***Table 1*** presents mean and median of chorological age, dental age and ALP concentration between all subjects. Twenty subjects were included and 50% of them were boys. The median chronological age of all the subjects was 9.75 years (8.08-13.42 years). The median Demirjian’s score for dental age was 92.45 (77.2 – 98.9). The median ALP concentration was 267.45 pg/mL (95.26 – 458.9 pg/mL).

**Table 1.** Mean and median of chorological age, dental age and ALP concentration between 20 subjects.

| | N | Mean ± SD | Median (lower-upper) |
| --- | --- | --- | --- |
| Chronological age (years) | 20 | 10.504 ± 1.895 | 9.75 ( 8.08 – 13.42 ) |
| Dental age (Demirjian's score) | 20 | 91.74 ± 5.972 | 92.45 ( 77.2 – 98.9 ) |
| ALP concentration (pg/mL) | 20 | 291.563 ± 115.135 | 267.45 ( 95.26 – 458.9 ) |

Out of 20 subjects, data were analyzed separately for girls, boys, and for both sex together. The mean biological age was 10.127 years for girls, 10.943 years for boys, and 10.504 years for the whole study group. ***Table 2*** showed mean of dental age and ALP concentrations according to sex and chronological age. Chronological age are divided by median value (11 years), in which 13 subjects belong to group 1 (<11 years old) and 7 subjects belong to group 2 (≥11 years). Sex proportion did not show significant correlation with neither dental age nor ALP concentration.

**Table 2.**
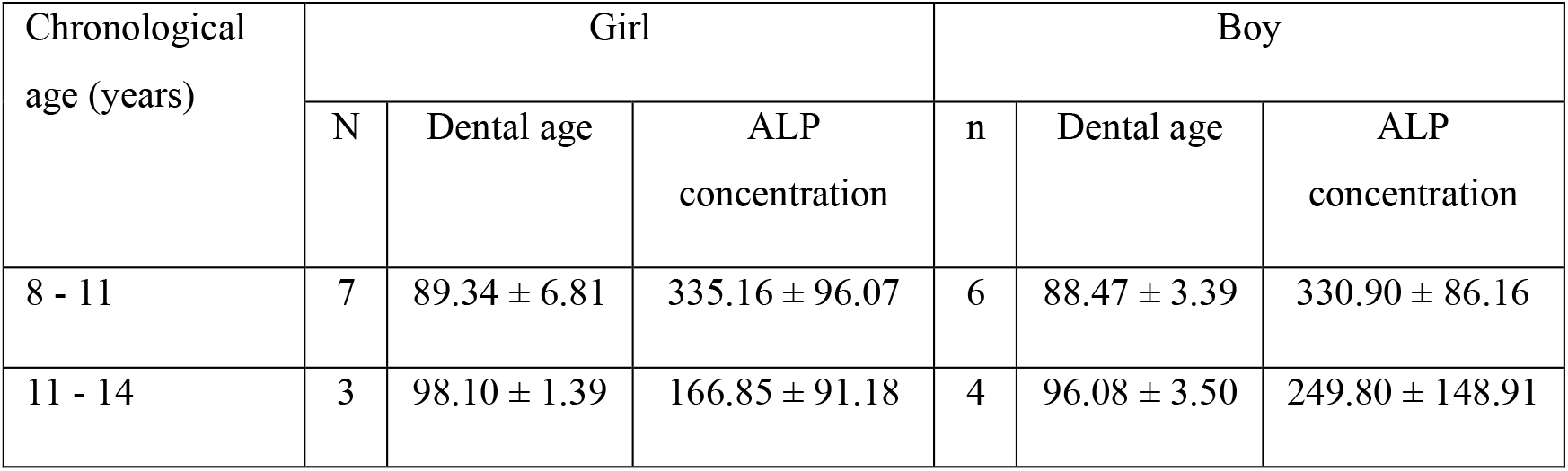
Mean of dental age and ALP concentrations according to sex and chronological age.

| Chronological<br>age (years) | Girl |  |  | Boy |  |  |
| --- | --- | --- | --- | --- | --- | --- |
|  | N | Dental age | ALP<br>concentration | n | Dental age | ALP<br>concentration |
| 8 - 11 | 7 | 89.34 ± 6.81 | 335.16 ± 96.07 | 6 | 88.47 ± 3.39 | 330.90 ± 86.16 |
| 11 - 14 | 3 | 98.10 ± 1.39 | 166.85 ± 91.18 | 4 | 96.08 ± 3.50 | 249.80 ± 148.91 |

We estimated each variable’snormality distributional assumptions using Kolmogorov-Smirnov and Shapiro-Wilk test. Normality test for the main variables can be seen in ***Table 3***. Statistical analysis with Kolmogorov-Smirnov and Shapiro-Wilk revealed that dental age and ALP concentrations are normally distributed, whereas chronological age isn’t normally distributed.

**Table 3.**
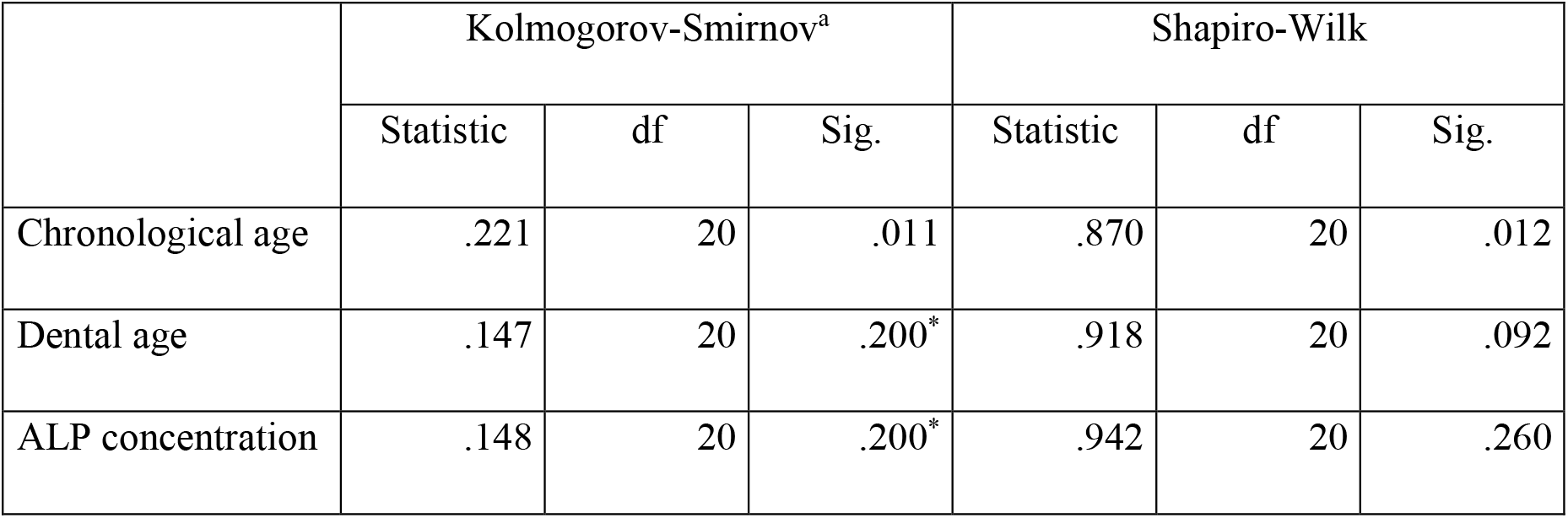
Kolmogorov-Smirnov and Shapiro-Wilk normality tests for main variables.

### Relationship between dental age and ALP concentration

The results of the correlational analysis between dental age (Demirjian’s score) and ALP concentration are summarized in ***Table 4***. Among general characteristics, ALP concentration showed a significantly negative correlation with dental age (*r*=−0.780; *p*<0.001).

**Table 4.** Correlation between dental age and ALP concentration.

| Variable |  | dental age | ALP<br>concentration |
| --- | --- | --- | --- |
| dental age | Correlation Coefficient | 1.000 | -.780** |
|  | Sig. (2-tailed) | . | .000 |
|  | N | 20 | 20 |
| ALP concentration | Correlation Coefficient | -.780** | 1.000 |
|  | Sig. (2-tailed) | .000 | . |
|  | N | 20 | 20 |

### Relationship between dental age and chronological age

The results of the correlational analysis between dental age (Demirjian’s score) and chronological age are summarized in ***Table 5***. Among general characteristics, chronological age showed a very strong positive correlation with dental age (*r*=+0.804; *p*<0.001).

**Table 5.** Correlation between dental age and chronological age.

| Variable |  |  | dental age | chronological age |
| --- | --- | --- | --- | --- |
| Spearman's rho | dental age | Correlation Coefficient | 1.000 | .804** |
|  |  | Sig. (2-tailed) | . | .000 |
|  |  | N | 20 | 20 |
|  | chronological age | Correlation Coefficient | .804** | 1.000 |
|  |  | Sig. (2-tailed) | .000 | . |
|  |  | N | 20 | 20 |
| **. Correlation is significant at the 0.01 level (2-tailed). |  |  |  |  |

### Relationship between chronological age and ALP concentration

The results of the correlational analysis between chronological age and ALP concentration are summarized in ***Table 6***. Among general characteristics, ALP concentration showed a moderate negative correlation with dental age (*r*=−0.508; *p*=0.022).

**Table 6.** Correlation between chronological age and ALP concentration.

|  |  |  | chronological age | ALP concentration |
| --- | --- | --- | --- | --- |
| Spearman's rho | chronological age | Correlation Coefficient | 1.000 | -.508* |
|  |  | Sig. (2-tailed) | . | .022 |
|  |  | N | 20 | 20 |
|  | ALP concentration | Correlation Coefficient | -.508* | 1.000 |
|  |  | Sig. (2-tailed) | .022 | . |
|  |  | N | 20 | 20 |
| *. Correlation is significant at the 0.05 level (2-tailed). |  |  |  |  |

## DISCUSSION

Various methods of dental age estimation are proposed by Demirjian, Nolla, Willems and Haavikko for growing individuals. All methods to estimate dental age are based on the following characteristics: eruption of the permanent tooth, level of root resorption of the primary tooth, and state of development of the root of the permanent teeth. Knowledge of deciduous crown formation times is useful in forensic anthropology and when aging juvenile remains from an archaeological context^38^. The findings of the present study showed that the Demirjian method had great accuracy for determining dental age in Indonesian healthy children in both sex. From the findings of the presented studies, a lack of precision and reliability between values of the dental age and salivary ALP to chronological age occurs in older age, because complete mineralization of the tooth root were increased, due to great variations of food intake and development in the difference socio-economic status^39-40^.

The results obtained in this cross-sectional study also demonstrated that the salivary ALP values were significantly correlated to biological age (Demirjian’s score) and chronological age. The values obtained in the study showed a high range of standard deviation which could have been because of great individual variation with regard to food intake and recent development. Salivary ALP has a vital role that may account for qualitative and quantitative dental maturation and remineralization. In contrast to serum bone ALP concentrations, salivary ALP concentrations are rarely used in routine biochemical tests^41^. Although salivary ALP concentrations can be performed in almost all laboratories, most pediatric dentists and forensic odontologists are doubted that its value can be altered by pathological conditions in dental tissue. Rise in salivary ALP levels can reflect activity of osteoblasts in spite of inflammation and destruction of healthy tissues such as dental caries or bone fracture^42^. Keeping in mind it was hypothesized that both physiologic and pathologic conditions have been associated with rise in serum ALP levels, possibly like bone growth, bone pathologies, hepatobiliary diseases and diabetes mellitus might have been associated with rise in salivary ALP levels too^43^.

### Strength and limitation

In the current study, we explored the relationship between chronological age and various indexes indicating dental maturation. One of the most significant findings to emerge from this study is that dental age (Demirjian’s score) and ALP concentration had a very strong correlation with chronological age and didn’t affect by sex bias. However, as noted before, discrepancies between chronological and biological ages are greater with increasing chronological age, despite the fact that advanced ages are associated with increased physical health problems such as dental caries, gingivitis and chronic periodontitis. With increasing age, the number of valid and meaningful dental characteristics diminishes, and therefore the accuracy of dental age assessment decreases^44^. In forensic dentistry, an accepted range of error values between the estimated dental age and chronological age for children is between 0.5-1.0 year^45^. According to the findings of the present study, Demirjian’s method and salivary ALP met these criteria to estimate biological age in the majority of Indonesian children. Another limitation of our study is the lack number of subjects, which can produce false-positive results, difficult statistical analysis or over-estimate the magnitude of an association between chronological age, dental age and salivary ALP.

## CONCLUSION

On the basis of the results of this study, it can be concluded that there is a strong correlation between dental age, chronological age, and salivary alkaline phosphatase among Indonesian children. We emphasize that dental age and salivary alkaline phosphatase have been developed to estimate the same, but accuracy of these methods are defined by their ability to arrive at an age as close to the chronological age, within acceptable error limits. The most important things are to combine several methods to improve the accuracy, reproducibility, precision and to reduce the discrepancy of actual age. Each technique should be feasible, simple and has a convenient approach for subjects.

## Data Availability

The datasets generated during and/or analyzed during the current study are not publicly available due to protecting participant confidentiality but are available from the corresponding author on reasonable request.

http://repository.unair.ac.id/82247/

## AVAILABILITY OF DATA AND MATERIAL

### FINANCIAL SUPPORT AND SPONSORSHIP

Nil.

## APPENDICES

